# Robust, reproducible clinical patterns in hospitalised patients with COVID-19

**DOI:** 10.1101/2020.08.14.20168088

**Authors:** Jonathan E Millar, Lucile Neyton, Sohan Seth, Jake Dunning, Laura Merson, Srin Murthy, Clark D Russell, Sean Keating, Maaike Swets, Carole Sudre, Timothy Spector, Sebastien Ourselin, Claire Steves, Jonathan Wolf, ISARIC-4C Investigators, Annemarie Docherty, Ewen Harrison, Peter Openshaw, Calum Semple, J. Kenneth Baillie

## Abstract

**Background:** Severe COVID-19 is characterised by fever, cough, and dyspnoea. Symptoms affecting other organ systems have been reported. However, it is the clinical associations of different patterns of symptoms which influence diagnostic and therapeutic decision-making. In this study, we applied simple machine learning techniques to a large prospective cohort of hospitalised patients with COVID-19 identify clinically meaningful sub-groups.

**Methods:** We obtained structured clinical data on 59 011 patients in the UK (the ISARIC Coronavirus Clinical Characterisation Consortium, 4C) and used a principled, unsupervised clustering approach to partition the first 25 477 cases according to symptoms reported at recruitment. We validated our findings in a second group of 33 534 cases recruited to ISARIC-4C, and in 4 445 cases recruited to a separate study of community cases.

**Findings:** Unsupervised clustering identified distinct sub-groups. First, a core symptom set of fever, cough, and dyspnoea, which co-occurred with additional symptoms in three further patterns: fatigue and confusion, diarrhoea and vomiting, or productive cough. Presentations with a single reported symptom of dyspnoea or confusion were common, and a subgroup of patients reported few or no symptoms. Patients presenting with gastrointestinal symptoms were more commonly female, had a longer duration of symptoms before presentation, and had lower 30-day mortality. Patients presenting with confusion, with or without core symptoms, were older and had a higher unadjusted mortality. Symptom clusters were highly consistent in replication analysis using a further 35446 individuals subsequently recruited to ISARIC-4C. Similar patterns were externally verified in 4445 patients from a study of self-reported symptoms of mild disease.

**Interpretation:** The large scale of the ISARIC-4C study enabled robust, granular discovery and replication of patient clusters. Clinical interpretation is necessary to determine which of these observations have practical utility. We propose that four patterns are usefully distinct from the core symptom groups: gastro-intestinal disease, productive cough, confusion, and pauci-symptomatic presentations. Importantly, each is associated with an in-hospital mortality which differs from that of patients with core symptoms. These observations deepen our understanding of COVID-19 and will influence clinical diagnosis, risk prediction, and future mechanistic and clinical studies.

**Funding:** Medical Research Council; National Institute Health Research; Wellcome Trust; Department for International Development; Bill and Melinda Gates Foundation; Liverpool Experimental Cancer Medicine Centre.

## Research in context

### Evidence before this study

We searched PubMed for articles published between 1st January and 31st August, 2020, using the search terms “COVID-19” OR “SARS-CoV-2” OR “2019-nCoV” AND “Symptom” OR “Feature” or “Characteristic”. No language restriction was enforced. An unprecedented number of articles have been published on COVID-19 since the beginning of the pandemic including many observational studies from a range of patient populations. Most are of limited size or scope. A report from the WHO-China Joint Mission provided an early description of COVID-19 symptoms in a large population of community and hospital cases and established fever, cough, and dyspnoea as cardinal symptoms. A variety of other symptoms have been reported and have been the subject of focused investigation, in particular gastrointestinal and neurological manifestations. Our work in mild cases identifies several distinct sub-groups with varied outcomes using time series data from 2700 community cases. No study has reported systematic symptom patterns in severe or fatal Covid-19.

### Added value of this study

We applied unsupervised machine learning techniques to symptom data from three cohorts of patients with COVID-19, totalling 63,456 individuals. We find that among hospitalised patients, symptoms occur in distinct groupings. Four clinically-relevant patterns are distinct from the core symptom group of cough, fever and dyspnoea. Subgroups of patients with gastro-intestinal disease, productive cough, confusion, and pauci-symptomatic presentations were found, each of which has a distinct association with baseline characteristics and outcome.

### Implications of all the available evidence

Our study is a comprehensive analysis of symptom patterns in hospitalised patients with Covid-19, of unprecedented scale and granularity. Our results are systematically validated in test set and in an external dataset. The prevalence of each symptom is consistent with previous reports, but we observe that these symptoms occur together in distinct and consistent groupings associated with different probabilities of death. This deepens our understanding of this new disease and is important for diagnosis, risk prediction, and the design of future clinical trials.

## Introduction

Severe coronavirus-19 disease (COVID-19), that is, confirmed COVID-19 as the primary reason for hospitalisation, is characterised by a triad of symptoms: cough, fever and dyspnoea. However it is clear that COVID-19 is not a homogeneous clinical entity. Important biological differences are likely to exist between patient subgroups, as is seen in other forms of critical illness including sepsis,^1^ pancreatitis,^2^, and dengue.^3^ Remarkably, this is already evident for COVID-19: highly significant sub-group effects were seen in the first drug trial to demonstrate an improvement in mortality, dexamethasone.^4^ The recognition of clinical similarities between patients is a fundamental unit of medical progress. Grouping patients enables us to select appropriate diagnostic tests, predict response to therapy, and to prognosticate. Simple machine learning methods can reveal patterns in clinical symptoms^5^ with diagnostic and therapeutic relevance.^6^

We employed unsupervised machine learning techniques in a large, prospective cohort of hospitalised patients with confirmed infection with SARS-CoV-2, to characterise the symptoms of severe COVID-19 and to identify clinical subgroups.

## Results

For the initial cohort, 33 468 patients were enrolled to the ISARIC-4C study, of which 7 991 had fully missing symptom data and were excluded from our analysis (Supplementary Figure S1). The baseline characteristics of included patients (25 477) are detailed in Supplementary Table 1. Overall, the median age was 73 (IQR 59-83) years and the majority of patients were male (15 046, 59%). On average, individuals presented to hospital 4 (IQR 1-8) days after the onset of symptoms.

### Symptom prevalence and relationships

Cough was the most prevalent symptom (68% (95%CI 67.5-68.6%)), followed by fever (66.4% (95%CI 65.8-67%)), and dyspnoea (65.2% (95%CI 64.6-65.8%)) (fig. 1 a and Supplementary Table 2). Furthermore, these were the only symptoms to be reported by greater than half of participants. The prevalence of individual symptoms varied with age (Supplementary Figure S2). Fever was less marked at the extremes of age, an observation which was also evident for dyspnoea, and, with the exception of those aged >90 years, for cough. Similarly, rash and runny nose were limited mostly to those aged <20 years, especially to those aged under 10 years. In sum, there were 4 335 unique symptom combinations in the cohort, the most frequent being fever, cough, and dyspnoea (1430, 5.6%)(fig. 1 a).

**Figure 1:**
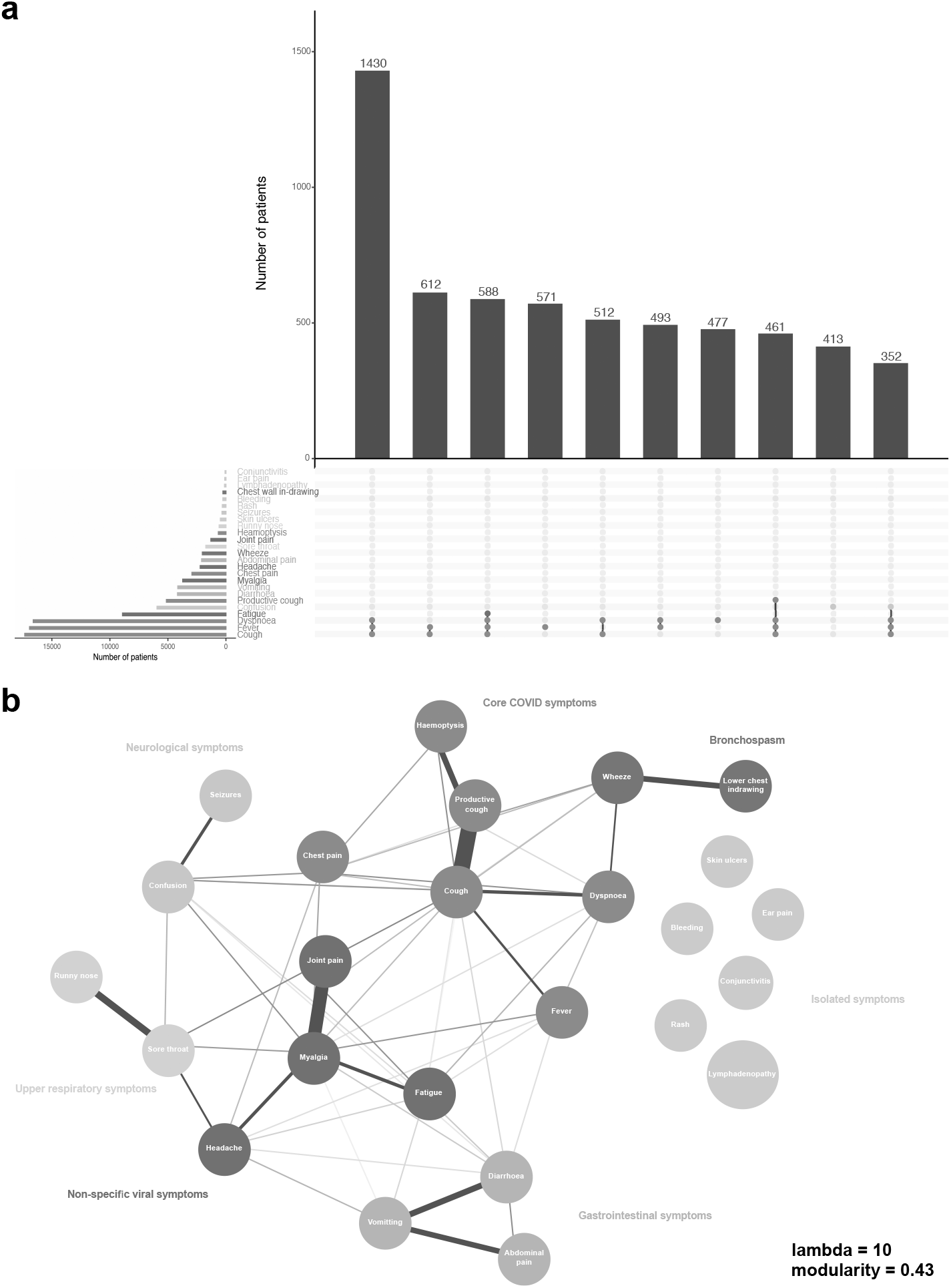
Symptom prevalence and relationships. a. Symptom prevalence. Upset plot, intersections describe the ‘top 10’ symptom combinations within the cohort. The upper graph charts the total number of patients exhibiting these symptom sets. The lower graph charts the total number of patients with each symptom in the cohort. b. Symptom network graph, derived using eLasso. Lines between symptom nodes illustrate conditional dependencies. A thicker line width and darker hue represents a stronger positive conditional dependence. Red lines represent a negative conditional dependence.

To explore the relationships between symptoms, we fitted an Ising model, employing L1-regularised logistic regression. The majority of symptoms exhibited some degree of conditional dependence with at least one other, however there were several that occurred independently; skin ulcers, rash, bleeding, lymphadenopathy, ear pain, and conjunctivitis (fig. 1 b). All of which had a low prevalence (<5%). Uniquely, confusion was negatively associated with cough, myalgia, sore throat, and diarrhoea. Groupings of symptoms with interconnected, positive conditional dependencies were appreciable from inspection of the network graph. To formalise these structures, we used a short random walk algorithm. Excluding the 6 orphan symptoms, 6 distinct communities were identified (modularity=0.43). These include: core COVID-19 (fever, cough, and dyspnoea), upper respiratory, bronchospasm, gastrointestinal (GI), neurological, and non-specific viral symptom sets (fig. 1 b).

### Patient clusters by symptom pattern

To identify symptom groupings within the study cohort, we performed unsupervised partitional clustering. An *a priori* assessment suggested that 7 clusters were optimal. This combined the inflection points in the decline in total sum of squares and the rise in gap statistic, with the nearest peak in average silhouette width (Supplementary Figure S3).

The patterns of symptoms within the seven clusters are shown in fig. 2 a. Based on the cluster medoid case, we characterised them as: core COVID-19 symptoms (fever, cough, and dyspnoea); core symptoms plus fatigue and confusion; productive cough; gastrointestinal (GI) symptoms; pauci-symptomatic; afebrile; and confusion. The core symptoms cluster accounted for the largest number of patients (9 364, 36.8%) and the GI symptoms cluster the fewest (1 327, 5.2%). Measures of cluster internal validity and stability are presented in Supplementary Figure S4 a.

**Figure 2:**
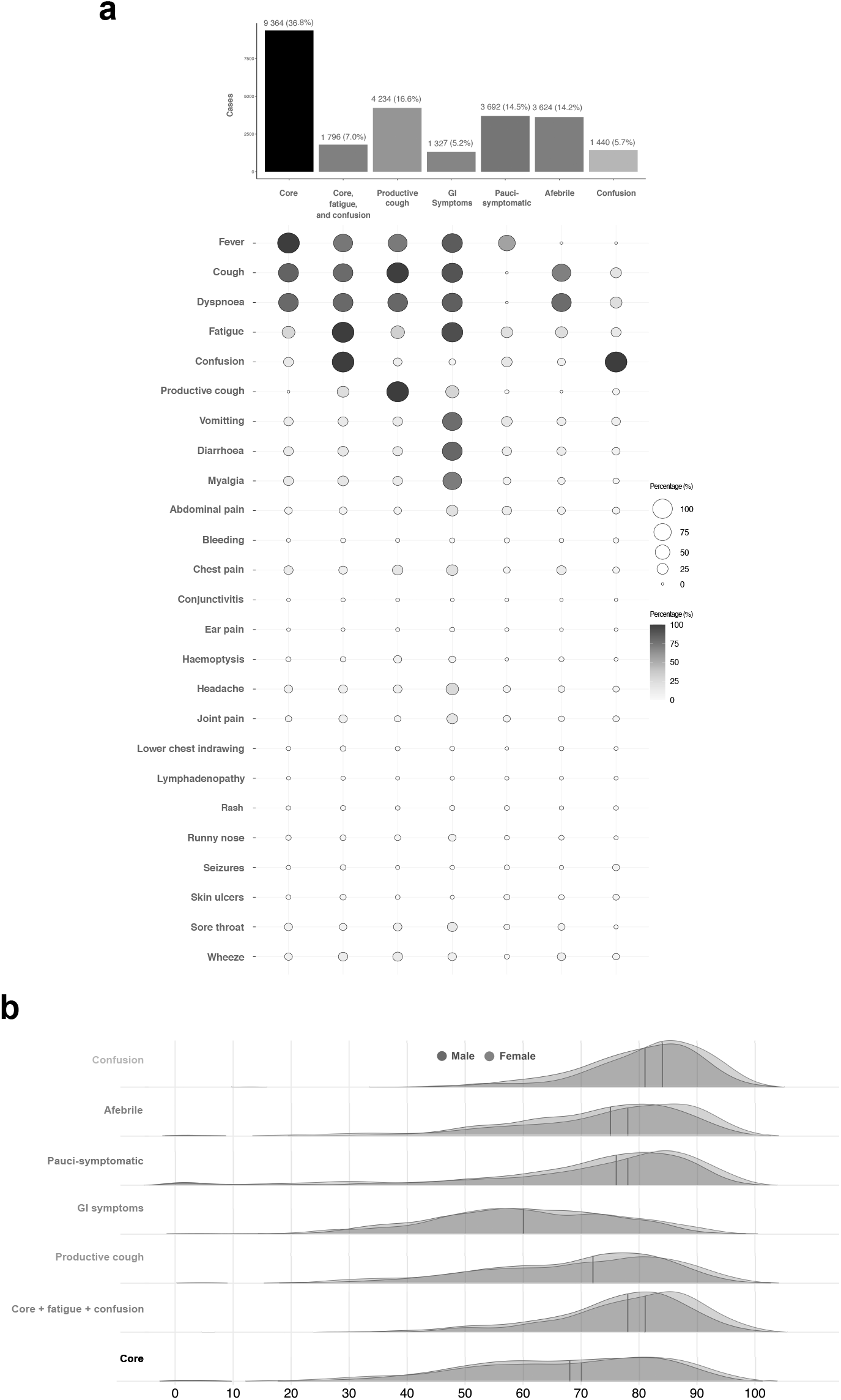
Symptom clusters. a. Cluster identities, proportions, and patterns. Data are presented as count (percentage) b. Distribution of age by symptom cluster. Density plots, solid lines represent the median age.

To examine the implications of our handling of missing data, we performed a sensitivity analysis by clustering only cases with fully complete symptom data (12 712, 49.9%). This analysis retained the cluster structure, with the exception of the afebrile cluster, in which the medoid case exhibited dyspnoea alone (Supplementary Figure S4 b). The simple agreement between iterations was 89.2%, with a Cohen’s kappa of 0.86 (p <0.001).

To validate the clusters identified by our primary analysis, we undertook two validation steps. First, we repeated our clustering approach on a cohort of patients enrolled to the ISARIC CCP-UK after those in the primary analysis and up until 7th July, 2020 (n=35 446). Of these, 1 912 (5.4%) had fully incomplete symptom data and were excluded from analysis. Clustering returned identical cluster medoids, with the exception of the afebrile cluster, in which cough was no longer implicated (Supplementary Figure S5). This cluster was also reduced in relative size (14.2% to 6.7%).

Secondly, our clustering approach was replicated in an outpatient cohort (COVID Symptom Study, n=4 445). This study records several overlapping or closely associated symptoms. Despite differences in study design and population, similar symptom groupings are discernible, including GI (cluster 1), pauci-symptomatic (cluster 2), and confusion clusters (cluster 5) (Supplementary Figure S6).

### Association of patient characteristics with symptom cluster

Compared to the cohort average (73 years (59-83)), those in the GI symptoms cluster were younger (60 years (49-72)), while those with core symptoms, fatigue and confusion (79 years (70-85)) or confusion alone (82 years (75-88)), were older (fig. 2 b). The GI symptom cluster also had the highest proportion of female patients (628, 47%). These differences were accompanied by variations in the median time from symptom onset to hospital admission between symptom clusters (p <0.001) (fig. 3 a), and similarly, in the burden of co-morbidity (fig. 3 b).

**Figure 3:**
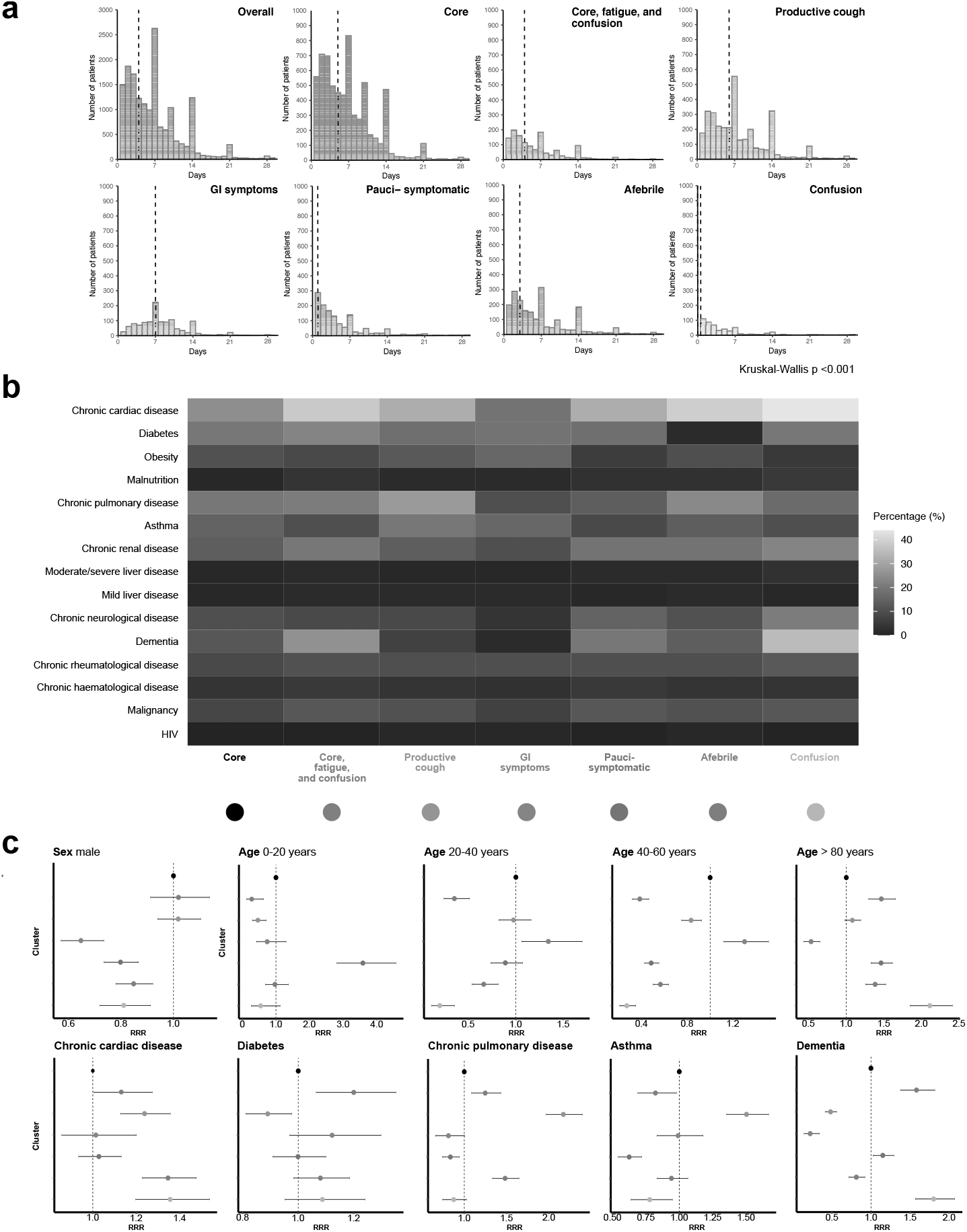
Patient characteristics and symptom clusters. a. Time from symptom onset to hospital admission. Data are presented as counts in single day bins. Vertical dashed lines represent the median time (days). b. Co-morbidities by symptom cluster. Percentage of individuals with co-morbidity at time of admission. c. Association between patient characteristics and symptom cluster membership. Multinomial regression, presented as odds ratio (95% confidence interval). Core symptoms chosen as reference cluster. The age group 60-80 years serves as the reference group for age.

To quantify these differences we used a multinomial logistic regression model. Taking the largest cluster (core symptoms) as our reference, we included major demographic variables and co-morbidities in the analysis (fig. 3 c and Supplementary Table S3). By comparison, those in the GI symptoms (RRR 0.66, 95% CI 0.60 to 0.73, p<0.001), afebrile (RRR 0.84, 95%CI 0.76 to 0.92, p<0.001), confusion (RRR 0.79, 95%CI 0.70 to 0.90, p<0.001), and pauci-symptomatic (RRR 0.78, 95%CI 0.72 to 0.86, p<0.001) clusters, were more likely to be female (fig. 3 c). The association between advanced age and confusion was mirrored by a higher prevalence of dementia in these groups (fig. 3 c and Supplementary Figure S3 a). Patients assigned to the productive cough cluster were more likely to suffer from chronic pulmonary diseases (RRR 2.07, 95%CI 1.84 to 2.33, p<0.001) and asthma (RRR 1.56, 95% CI 1.37 to 1.77, p<0.001) (fig. 3 c).

### Association between symptom cluster and outcome

Overall, the unadjusted in-hospital mortality was 35%, with 24% having an incomplete hospital episode at the end of follow-up. Outcomes, stratified by cluster allocation, are detailed in Supplementary Table S1. To assess differences in mortality between symptom clusters, we first compared Kaplan-Meier curves (log-rank test, p<0.001) (fig. 4 a). For the largest cluster, core symptoms, unadjusted in-hospital mortality was 33%. The lowest mortality was found in the GI symptoms cluster (18%) and the highest in the core, fatigue, and confusion cluster (53%). Subsequently, we used a Cox proportional hazards model to account for the influence of age and sex (fig. 4 b). Those in the core, fatigue, and confusion cluster remained at the highest risk of death when compared to those with core symptoms, (HR 1.26, 95%CI 1.15-1.37, p <0.001). However, membership of the confusion cluster was no longer associated with an increased risk of death (HR 0.92, 95%CI 0.83-1.02, p=0.096). Those in the productive cough, GI, and pauci-symptomatic clusters continued to attract a lower risk of death (fig. 4 b). Given that there was evidence of variation in risk over time for some clusters, we performed Restricted Mean Survival Time (RMST) analyses (Supplementary Table S4). By this method, males in the core, fatigue, and confusion cluster had the poorest survival compared to core symptoms, mean survival difference at 30-days −1.9 days (95% CI −2.8 - 1.1).

**Figure 4:**
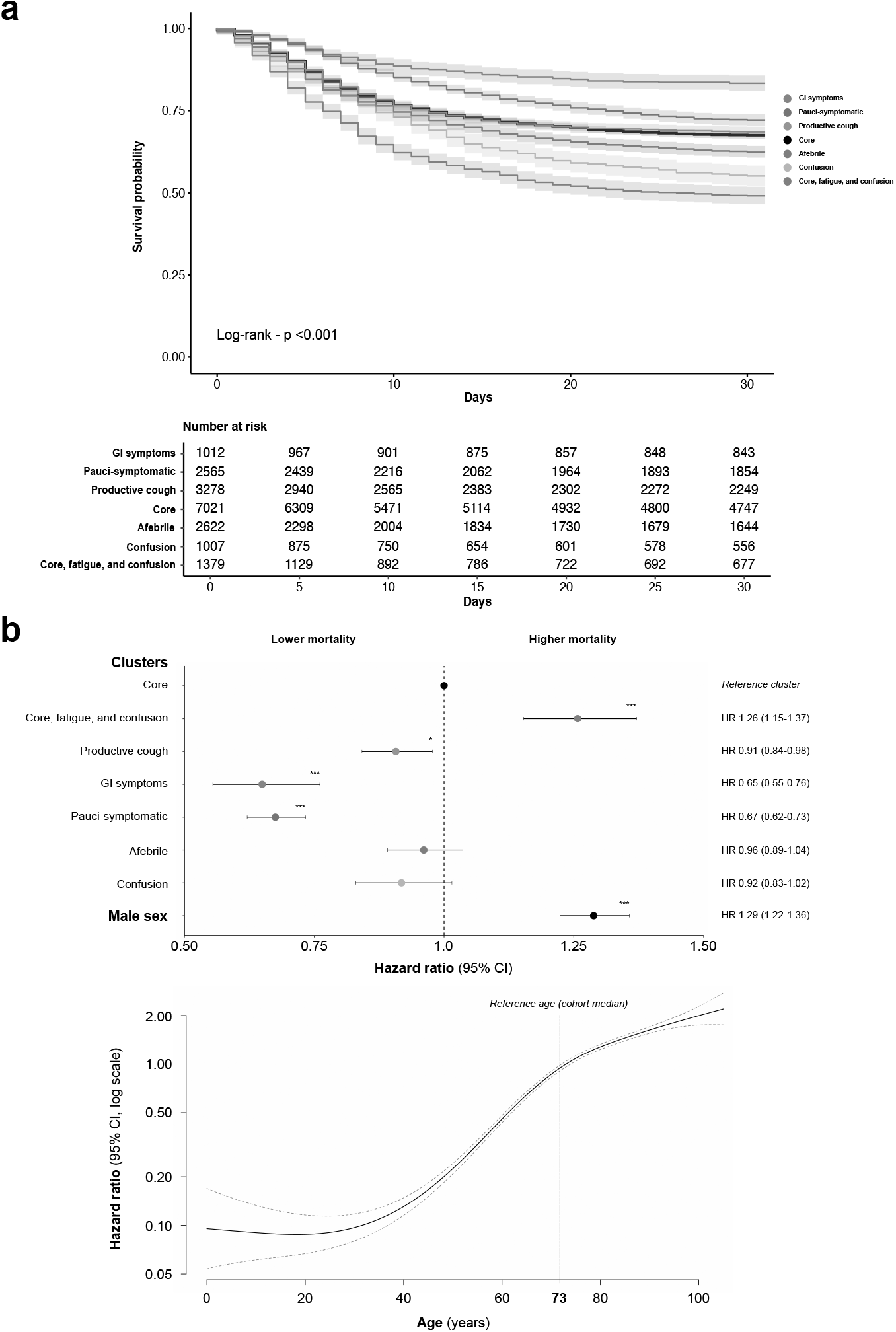
Symptom clusters and survival. a. Unadjusted 30-day in-hospital mortality by cluster. Kaplan-Meier curves, those discharged before 30 days were assumed to have survived until the end of follow-up. b. The risk of 30 day in-hospital mortality, adjusted for age and sex. Upper panel, Forest plot, showing results of a Cox proportional hazards model. Lower panel, Hazard ratio associated with varying age. Data are presented as hazard ratio (HR) and 95% confidence intervals. Red dotted line −95% CI Core symptoms serves as the reference symptom cluster. The cohort median age serves as the reference age.

## Discussion

This study identifies distinct symptom clusters in a large cohort of hospitalised patients with COVID-19. These clusters are internally robust and reproducible. To our knowledge, this report also provides the largest dataset of symptom prevalence in COVID-19 patients to date.

Knowledge of distinct symptom sub-phenotypes has potential importance for our mechanistic understanding of COVID-19. Two groups in our analysis have distinct clinical trajectories: those presenting with GI symptoms, and those presenting with confusion. Those in the GI cluster tended to be younger, more likely female, presented to hospital later, and had a higher probability of survival. Conversely, those with confusion (with or without fever, cough, and dyspnoea), were older, presented earlier, and had poorer outcomes. These data may be important for refining risk-prediction at the time of hospital admission. Similarly, the identification of a cluster in which patients had few symptoms other than confusion has implications for defining cases and for targeting testing, particularly in elderly patients. We suggest that patients presenting with isolated diarrhoea, vomiting, abdominal pain, or new confusion of unknown aetiology should be tested for COVID-19. Importantly, these clusters have divergent outcomes from those with core COVID-19 symptoms.

Gastrointestinal infection occurred in SARS,^7^ MERS,^7^ and was reported in early descriptions of COVID-19.^8^ Single-cell transcriptomic analysis from ileum and colon has demonstrated the ability of SARS-CoV-2 to infect enterocytes,^9^ leading to the possibility of an enteric form of COVID-19.^10^

Our identification of a group of COVID-19 patients presenting with confusion, and not other symptoms, has direct clinical implications. The causal mechanisms leading to this presentation may not be directly related to COVID-19: confusion is a common nonspecific presenting symptom, particularly among elderly patients. However, emerging evidence suggests that SARS-CoV-2 may enter the CNS directly via the olfactory bulb, and precipitates inflammation with direct effects on the brain.^11^ Imoprtantly, this group are at higher absolute risk of death.

These data may offer a starting point for predictive enrichment in clinical trials.^12^ Predictive enrichment based on symptom clustering is expected to perform best where causal relationships exist between fundamental biological or genomic features of disease and the clinical manifestations of severe illness.^13^ Similar relationships have been described in conditions as diverse as schizophrenia^14^ and asthma.^15^ The re-analysis of trial results based on patient clustering may reveal differential treatment effects.^16^

The pooled symptom prevalences reported in the largest meta-analysis of independent studies (including 24401 individuals), as well as those found in the WHO-China Joint Mission Report, are broadly consistent with the data reported here, taking into account the higher severity of illness and more advanced age in the ISARIC 4C cohort.^17,18^ However, the prevalence of GI symptoms and of confusion was higher in our study.

For survival analysis, statistical adjustment was limited to variables that we know to have been largely complete and not subject to significant confounding (age and sex). For some clusters, due to differences in disease trajectory, there was evidence of variation in risk of death with time. Although this violates the assumption of proportional hazards, RMST analyses confirmed the directions of effect. In handling missing symptom records, we sought to retain as much data as possible because missingness itself was informative. This approach was robust to a sensitivity analysis. For survival data, several methods of analysis were employed to reduce the risk of bias.

The ISARIC 4C study was limited to moderate to severe cases of COVID-19 presenting to hospital. Since symptoms were sought only at hospital admission, recall bias may affect symptom reporting. Additionally, a small number of symptoms now known to be associated with COVID-19, namely anosmia and ageusia,^19^ were not recorded. Therefore, our conclusions do not apply to individuals with mild disease.

Clustering a large, heterogeneous population requires investigators to strike a balance between parsimony and granularity. Each patient is unique, and there were 4335 symptom combinations in the ISARIC 4C cohort; hence, the most granular clustering would reveal 4335 distinct patterns of disease. Our purpose in grouping these patients is to reveal clinical patterns that will have practical utility. As we have argued previously, the question should not be “How many clusters exist?”, but rather, “Which clusters are potentially useful?.”^13^

This study of 68914 hospitalised patients with COVID-19 identified distinct symptom sub-groupings whose character and outcome differed significantly from those with the core symptoms of COVID-19. These observations deepen understanding of COVID-19 and will influence clinical diagnosis, risk prediction, and future mechanistic and clinical studies.

## Methods

### Study design, setting, and population

The ISARIC Cornavirus Clinical Characterisation Consortium (4C) study is an ongoing prospective cohort study, involving 260 acute hospital sites in England, Scotland, and Wales. The study builds on an international consensus protocol for investigation of new infectious diseases, the International Severe Acute Respiratory Infection Consortium/World Health Organisation Clinical Characterisation Protocol (ISARIC/WHO CCP),^20^ designed to enable internationally-harmonised clinical research during outbreaks.^21^ The protocol, revision history, case report form, information leaflets, consent forms, and details of the Independent Data and Material Access Committee are available at https://isaric4c.net. The UK study was approved by the South Central-Oxford C Research Ethics Committee (13/SC/0149) and by the Scotland A Research Ethics Committee (20/SS/0028). This study is reported in compliance with the STROBE guidelines.^22^

Patients included in the primary analysis were admitted to hospital between 6th February and 8th May, 2020. Inclusion criteria were all patients admitted to a participating hospital with laboratory proven or clinically highly suspected SARS-CoV-2 infection. Reverse transcription-PCR was the sole method of testing available during the study period. In the original CCP,^20^ the inclusion of patients with clinically-suspected infection reflects the design of this study as a preparedness protocol where laboratory tests may not be available, but in the context of this outbreak in the UK, site training emphasised the importance of enrolling only laboratory-confirmed cases. Patients who were admitted to hospital for an unrelated condition but who subsequently tested positive for SARS-CoV-2 were also included.

### Data collection

Data were collected on a case report form, developed by ISARIC and WHO in advance of this outbreak. From admission, data were uploaded to an electronic database (REDCap, Vanderbilt University, US; hosted by University of Oxford, UK). We recorded demographic details as well as patient co-morbidities, in-hospital clinical course, treatments, and outcomes.

The presence or absence of a pre-defined list of symptoms was assessed at hospital admission. These were: fever, cough, productive cough, haemoptysis, dyspnoea, wheeze, chest wall in-drawing, chest pain, fatigue, myalgia, joint pain, vomiting, diarrhoea, abdominal pain, headache, confusion, seizures, lymphadenopathy, ear pain, sore throat, runny nose, conjunctivitis, rash, and skin ulceration.

Similarly, the presence of pre-defined co-morbidities was recorded. These were: asthma, diabetes (type 1 and type 2), chronic cardiac disease, chronic haematological disease, chronic kidney disease, chronic neurological disease, chronic pulmonary disease (excluding asthma), dementia, HIV, malignancy, malnutrition, mild liver disease, moderate or severe liver disease, obesity, chronic rheumatological disease, and smoking history.

Outcomes data were collected for admission to critical care (Intensive Care Unit or High Dependency Unit), the use of invasive mechanical ventilation (IMV), and in-hospital mortality.

### Statistical analysis

Continuous data are summarised as median (inter-quartile range). Categorical data are summarized as frequency (percentage). Prevalence is reported as percentage (95% confidence interval (CI)). Confidence intervals were calculated for a binomial proportion using the Clopper-Pearson exact method. We analysed data using R (R Core Team, Version 4.0.0, Vienna, Austria). *p* values < 0.05 were deemed significant.

### Missing data

Given the extraordinary circumstances in which this study was conducted there was a large amount of missing data. No attempt at multiple imputation was made. In respect of symptom data, in many cases the presence of a positive symptom(s) was recorded, with the remainder missing. Exploration of the structure of missing symptom data (Supplementary Figure S1) suggests that this did not occur at random. In such cases, missing symptoms were recoded as being absent. Patients with fully missing data were excluded from the analysis.

#### Symptom network analysis

To explore the relationship between the 25 recorded symptoms we fitted an Ising model, using L1-regularised logistic regression with model selection by Extended Bayesian Information criteria (EBIC),^23^ using the R package *IsingFit*. A λ value of 10 was chosen to minimise spurious conditional dependencies. The partition of symptoms into communities was formalised using a short random walks method, with the R package *igraph*.^24^

#### Unsupervised partitional clustering

Symptom data for each patient, encoded as binary responses, were used to derive a Jaccard distance matrix. This was then supplied as the input to a k-medoids clustering algorithm. This is an unsupervised partitional algorithm that seeks to divide the sample into *k* clusters, where the arbitrary distance between any individual case and the case chosen as the centre of a cluster (medoid) is minimised. In this study we used a variation of this algorithm, Clustering for Large Applications (CLARA), with the R package *cluster*.^25^ CLARA, for the optimisation of computational runtime, performs iterations of k-medoids clustering on subsets of the data and selects the best performing result. We clustered 100 random sub-samples each consisting of approximately 10% of the analysed population (n=2500). Each sub-sample was used to select *k* medoid cases, after which every case in the dataset was assigned to the nearest medoid. The iteration in which the mean of the dissimilarities between cases and the nearest medoid was lowest was selected. Random sampling was performed deterministically to ensure consistency between our primary analysis and validation steps. Clustering was performed agnostic of patient demographics or outcome. The optimal number of clusters to specify to the algorithm was derived from a ‘majority’ assessment of three measures; total within sum of squares, average silhouette width, and gap statistic,^26^ in which a parsimonious solution was preferred. To assess the stability of clusters, we employed a non-parametric bootstrap-based strategy.^27^ This generated 1000 new datasets by randomly drawing samples from the initial dataset with replacement and applying the same clustering technique to each. Clustering results were then compared for each cluster identified in the primary analysis and the most similar cluster identified for each random re-sampling. A mean value for the Jaccard coefficient, for the sum of the comparisons, was generated for each cluster present in the primary analysis. As a sensitivity analysis, clustering was repeated on patients with only fully complete symptom data. Cluster allocations for individuals partitioned by both iterations were then compared using Cohen’s kappa and simple percentage agreement with the R package *irr*.

#### Clustering validation

We performed two validations, one internal and one external. Internally, we used symptom data for patients enrolled to ISARIC CCP-UK after those included in the primary analysis and until 7th July, 2020. These data were processed and analysed as for the primary cohort. Missing data were treated in the same fashion. Externally, our clustering strategy was replicated independently in a sub-sample of users from the COVID Symptom study app (developed by Zoe Global limited with input from scientists and clinicians from King’s College London and Massachusetts general hospital. Individuals with confirmed SARS-CoV-2 laboratory results, registering healthy on the app, with symptom duration of more than 7 days were included, considering the presentation at symptom peak to build the clustering.^28^

#### Multinomial regression modelling and time to admission

To quantify the relationship between demographic factors, co-morbidities and cluster membership we built a multinomial logistic regression model with the R package *nnet*. For binary variables, a missing value was assumed to correspond to the absence of a given co-morbidity. Individuals with missing values for age and sex were excluded from this analysis. The dependent variable was symptom cluster. The independent variables were; sex, age (categorical), chronic cardiac disease, chronic pulmonary disease, asthma, chronic kidney disease, chronic neurological disease, malignancy, chronic haematological disease, HIV, chronic rheumatological disease, dementia, malnutrition, chronic liver disease, and diabetes. Results are summarised as relative risk ratio (RRR) and 95% confidence intervals (95% CI). The average time from self-reported symptom onset to hospital admission in each cluster were compared using the Kruskal-Wallis test.

#### Survival analysis

To examine survival differences between symptom clusters we employed several methods. Kaplan-Meier curves for 30-day in-hospital mortality were calculated and compared using a log-rank test, with the R package *survival*. Individuals reported as being dead but with no outcome date were excluded. All individuals not reported as dead were presumed alive. Discharged individuals were retained within the at-risk set until the end of follow-up; thus, discharge did not compete death. Survival time was defined as the time in days between hospital admission and the reported outcome date. We then fitted a Cox proportional hazards model to the data with the *a priori* inclusion of age and sex as co-variates, using the R package *survival*. Given the non-linear effect of age on the risk of dath we fitted age with a penalised smoothing spline. Results are reported as hazard ratio (HR) and 95% CI. In anticipation that the risk of death per cluster varied over time, we also calculated restricted mean survival times at 30 days, insuring the analysis against violations of proportional hazards assumptions. This was performed with the R package *survRM2*. Results are reported as mean survival difference (days) and 95% CI.

## Data Availability

We welcome applications for data and material access through our Independent Data and Material Access Committee (https://isaric4c.net).

## Acknowledgements

The study protocol is available at http://isaric4c.net/protocols; study registry https://www.isrctn.com/ISRCTN66726260. This work uses data provided by patients and collected by the NHS as part of their care and support #DataSavesLives. We are extremely grateful to the frontline NHS clinical and research staff and volunteer medical students who collected these data in challenging circumstances; and the generosity of the patients and their families for their individual contributions in these difficult times. We also acknowledge the support of Jeremy J Farrar, Nahoko Shindo, Devika Dixit, Nipunie Rajapakse, Piero Olliaro, Lyndsey Castle, Martha Buckley, Debbie Malden, Katherine Newell, Kwame O’Neill, Emmanuelle Denis, Claire Petersen, Scott Mullaney, Sue MacFarlane, Chris Jones, Nicole Maziere, Katie Bullock, Emily Cass, William Reynolds, Milton Ashworth, Ben Catterall, Louise Cooper, Terry Foster, Paul Matthew Ridley, Anthony Evans, Catherine Hartley, Chris Dunn, D Sales, Diane Latawiec, Erwan Trochu, Eve Wilcock, Innocent Gerald Asiimwe, Isabel Garcia-Dorival, J Eunice Zhang, Jack Pilgrim, Jane A Armstrong, Jordan J Clark, Jordan Thomas, Katharine King, Katie Neville, Alexandra Ahmed, Krishanthi S Subramaniam, Lauren Lett, Laurence McEvoy, Libby van Tonder, Lucia Alicia Livoti, Nahida S Miah, Rebecca K Shears, Rebecca Louise Jensen, Rebekah Penrice-Randal, Robyn Kiy, Samantha Leanne Barlow, Shadia Khandaker, Soeren Metelmann, Tessa Prince, Trevor R Jones, Benjamin Brennan, Agnieska Szemiel, Siddharth Bakshi, Daniella Lefteri, Maria Mancini, Julien Martinez, Angela Elliott, Joyce Mitchell, John McLauchlan, Aislynn Taggart, Oslem Dincarslan, Annette Lake, Claire Petersen, Scott Mullaney, and Graham Cooke.

## ISARIC4C Investigators

Consortium Lead Investigator J Kenneth Baillie, Chief Investigator Malcolm G Semple, Co-Lead Investigator Peter JM Openshaw. ISARIC Clinical Coordinator Gail Carson. Co-Investigators: Beatrice Alex, Benjamin Bach, Wendy S Barclay, Debby Bogaert, Meera Chand, Graham S Cooke, Annemarie B Docherty, Jake Dunning, Ana da Silva Filipe, Tom Fletcher, Christopher A Green, Ewen M Harrison, Julian A Hiscox, Antonia Ying Wai Ho, Peter W Horby, Samreen Ijaz, Saye Khoo, Paul Klenerman, Andrew Law, Wei Shen Lim, Alexander, J Mentzer, Laura Merson, Alison M Meynert, Mahdad Noursadeghi, Shona C Moore, Massimo Palmarini, William A Paxton, Georgios Pollakis, Nicholas Price, Andrew Rambaut, David L Robertson, Clark D Russell, Vanessa Sancho-Shimizu, Janet T Scott, Louise Sigfrid, Tom Solomon, Shiranee Sriskandan, David Stuart, Charlotte Summers, Richard S Tedder, Emma C Thomson, Ryan S Thwaites, Lance CW Turtle, Maria Zambon. Project Managers Hayley Hardwick, Chloe Donohue, Jane Ewins, Wilna Oosthuyzen, Fiona Griffiths. Data Analysts: Lisa Norman, Riinu Pius, Tom M Drake, Cameron J Fairfield, Stephen Knight, Kenneth A Mclean, Derek Murphy, Catherine A Shaw. Data and Information System Manager: Jo Dalton, Michelle Girvan, Egle Saviciute, Stephanie Roberts Janet Harrison, Laura Marsh, Marie Connor. Data integration and presentation: Gary Leeming, Andrew Law, Ross Hendry. Material Management: William Greenhalf, Victoria Shaw, Sarah McDonald. Local Principal Investigators: Kayode Adeniji, Daniel Agranoff, Ken Agwuh, Dhiraj Ail, Ana Alegria, Brian Angus, Abdul Ashish, Dougal Atkinson, Shahedal Bari, Gavin Barlow, Stella Barnass, Nicholas Barrett, Christopher Bassford, David Baxter, Michael Beadsworth, Jolanta Bernatoniene, John Berridge, Nicola Best, Pieter Bothma, David Brealey, Robin Brittain-Long, Naomi Bulteel, Tom Burden, Andrew Burtenshaw, Vikki Caruth, David Chadwick, Duncan Chambler, Nigel Chee, Jenny Child, Srikanth Chukkambotla, Tom Clark, Paul Collini, Graham Cooke, Catherine Cosgrove, Jason Cupitt, Maria-Teresa Cutino-Moguel, Paul Dark, Chris Dawson, Samir Dervisevic, Phil Donnison, Sam Douthwaite, Ingrid DuRand, Ahilanadan Dushianthan, Tristan Dyer, Cariad Evans, Chi Eziefula, Chrisopher Fegan, Adam Finn, Duncan Fullerton, Sanjeev Garg, Sanjeev Garg, Atul Garg, Jo Godden, Arthur Goldsmith, Clive Graham, Elaine Hardy, Stuart Hartshorn, Daniel Harvey, Peter Havalda, Daniel B Hawcutt, Maria Hobrok, Luke Hodgson, Anita Holme, Anil Hormis, Michael Jacobs, Susan Jain, Paul Jennings, Agilan Kaliappan, Vidya Kasipandian, Stephen Kegg, Michael Kelsey, Jason Kendall, Caroline Kerrison, Ian Kerslake, Oliver Koch, Gouri Koduri, George Koshy, Shondipon Laha, Susan Larkin, Tamas Leiner, Patrick Lillie, James Limb, Vanessa Linnett, Jeff Little, Michael MacMahon, Emily MacNaughton, Ravish Mankregod, Huw Masson, Elijah Matovu, Katherine McCullough, Ruth McEwen, Manjula Meda, Gary Mills, Jane Minton, Mariyam Mirfenderesky, Kavya Mohandas, Quen Mok, James Moon, Elinoor Moore, Patrick Morgan, Craig Morris, Katherine Mortimore, Samuel Moses, Mbiye Mpenge, Rohinton Mulla, Michael Murphy, Megan Nagel, Thapas Nagarajan, Mark Nelson, Igor Otahal, Mark Pais, Selva Panchatsharam, Hassan Paraiso, Brij Patel, Justin Pepperell, Mark Peters, Mandeep Phull, Stefania Pintus, Jagtur Singh Pooni, Frank Post, David Price, Rachel Prout, Nikolas Rae, Henrik Reschreiter, Tim Reynolds, Neil Richardson, Mark Roberts, Devender Roberts, Alistair Rose, Guy Rousseau, Brendan Ryan, Taranprit Saluja, Aarti Shah, Prad Shanmuga, Anil Sharma, Anna Shawcross, Jeremy Sizer, Richard Smith, Catherine Snelson, Nick Spittle, Nikki Staines, Tom Stambach, Richard Stewart, Pradeep Subudhi, Tamas Szakmany, Kate Tatham, Jo Thomas, Chris Thompson, Robert Thompson, Ascanio Tridente, Darell Tupper-Carey, Mary Twagira, Andrew Ustianowski, Nick Vallotton, Lisa Vincent-Smith, Shico Visuvanathan, Alan Vuylsteke, Sam Waddy, Rachel Wake, Andrew Walden, Ingeborg Welters, Tony Whitehouse, Paul Whittaker, Ashley Whittington, Meme Wijesinghe, Martin Williams, Lawrence Wilson, Sarah Wilson, Stephen Winchester, Martin Wiselka, Adam Wolverson, Daniel G Wooton, Andrew Workman, Bryan Yates, Peter Young.

## Data sharing

We welcome applications for data and material access through our Independent Data and Material Access Committee (https://isaric4c.net). The lead author (the manuscript’s guarantor) affirms that the manuscript is an honest, accurate, and transparent account of the study being reported; that no important aspects of the study have been omitted; and that any discrepancies from the study as planned (and, if relevant, registered) have been explained. Dissemination ISARIC4C has a public facing website and twitter account **???**. We are engaging with print and internet press, television, radio, news, and documentary programme makers. The Corresponding Author has the right to grant on behalf of all authors and does grant on behalf of all authors, a worldwide licence (http://www.bmj.com/sites/default/files/BMJ%20Author%20Licence%20March%202013.doc) to the Publishers and its licensees in perpetuity, in all forms, formats and media (whether known now or created in the future), to i) publish, reproduce, distribute, display and store the Contribution, ii) translate the Contribution into other languages, create adaptations, reprints, include within collections and create summaries, extracts and/or, abstracts of the Contribution and convert or allow conversion into any format including without limitation audio, iii) create any other derivative work(s) based in whole or part on the on the Contribution, iv) to exploit all subsidiary rights to exploit all subsidiary rights that currently exist or as may exist in the future in the Contribution, v) the inclusion of electronic links from the Contribution to third party material where-ever it may be located; and, vi) licence any third party to do any or all of the above. All research articles will be made available on an open access basis (with authors being asked to pay an open access fee—see http://www.bmj.com/about-bmj/resources-authors/forms-policies-andchecklists/copyright-open-access-and-permission-reuse). The terms of such open access shall be governed by a Creative Commons licence—details as to which Creative Commons licence will apply to the research article are set out in our worldwide licence referred to above.

---

**Supplementary Figure 1.**
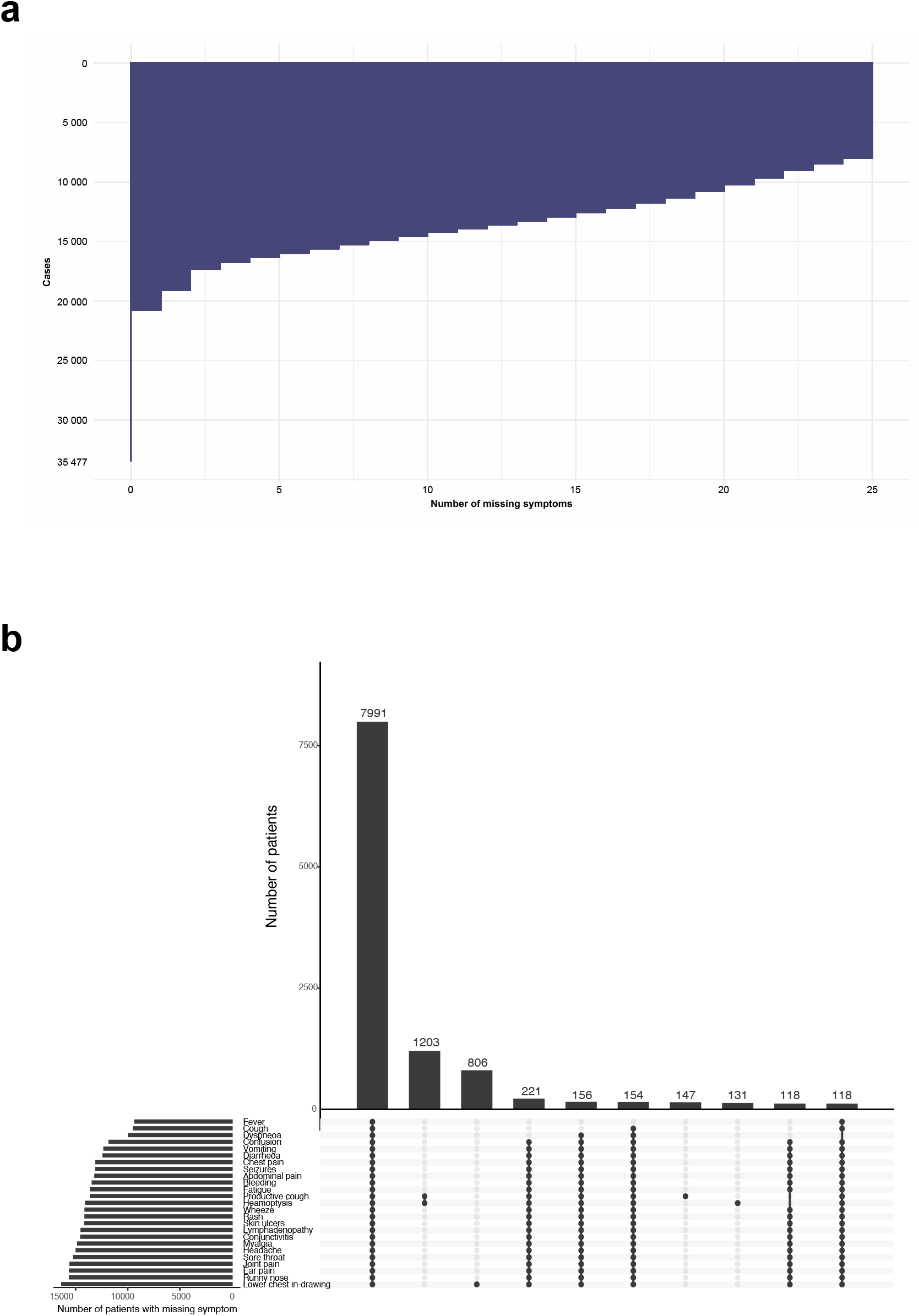
Missing symptom data. **a**. Summary of missing symptom data by case. **b**. Patterns of missing symptom data. Upset plot, intersections represent the ‘top 10’ missing symptom combinations. Upper graph summarises the number of patients with each pattern. Lower graph shows the total number of patients with missing data for each symptom.

**Supplementary Figure S2.**
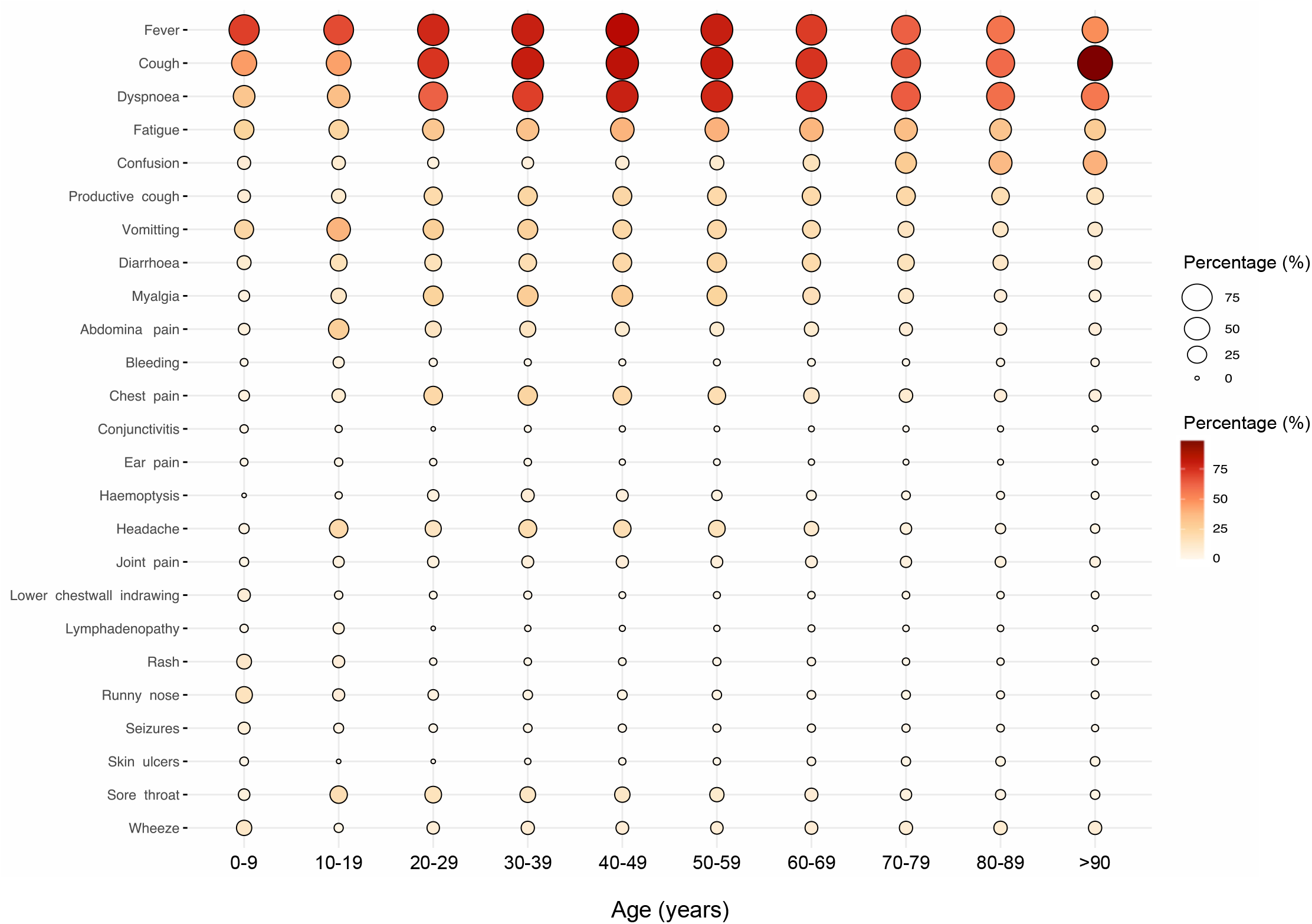
Symptom proportions and patterns by age. Data are presented as percentage (%) of patients exhibiting each symptom within each decile.

**Supplementary Figure 3.**
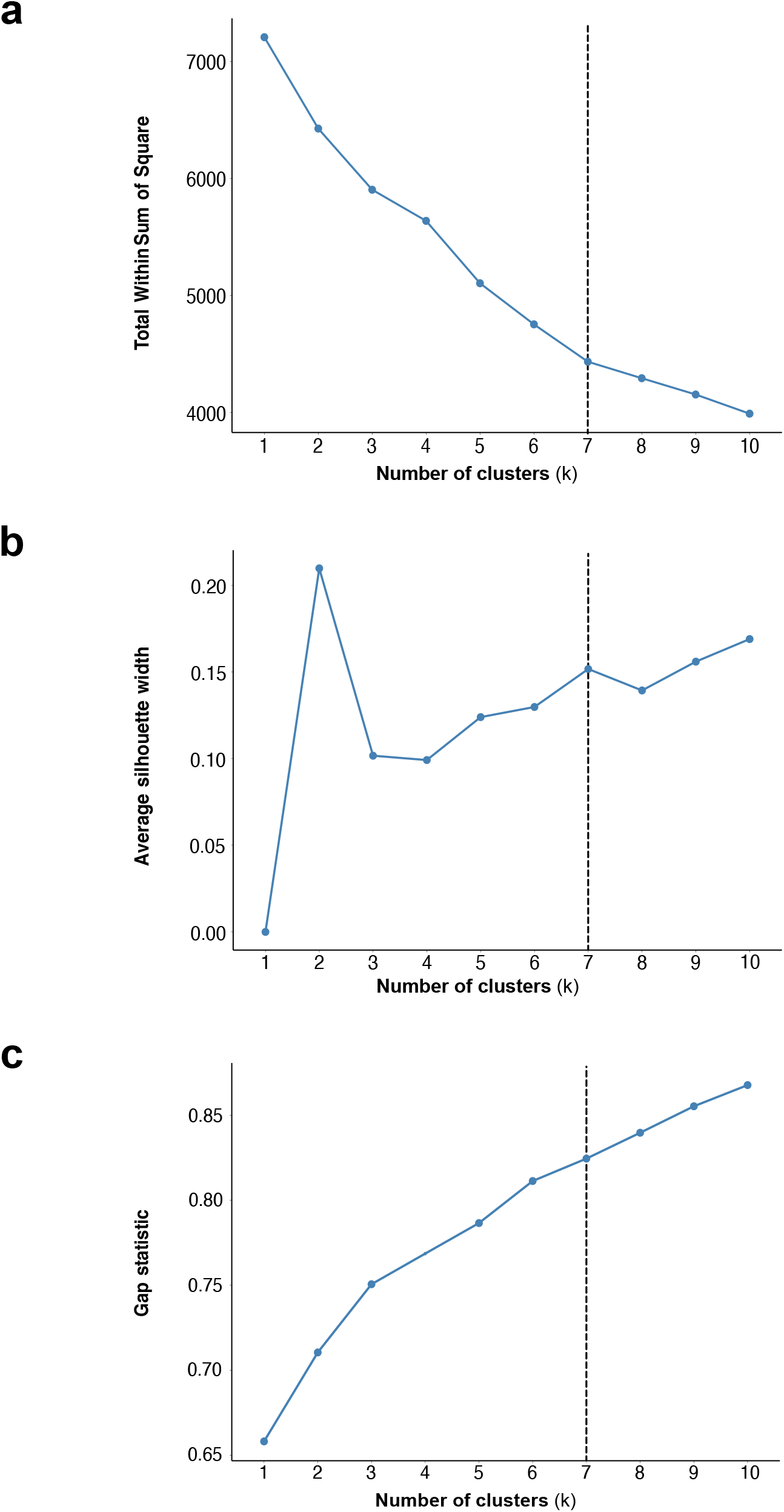
Measures of optimal cluster number. **a**. Total within sum of squares. b. Average silhouette width. c. Gap statistic. Data are presented across a range of solutions from k=1 to k=10. Vertical dashed line represents the chosen solution.

**Supplementary Figure 4.**
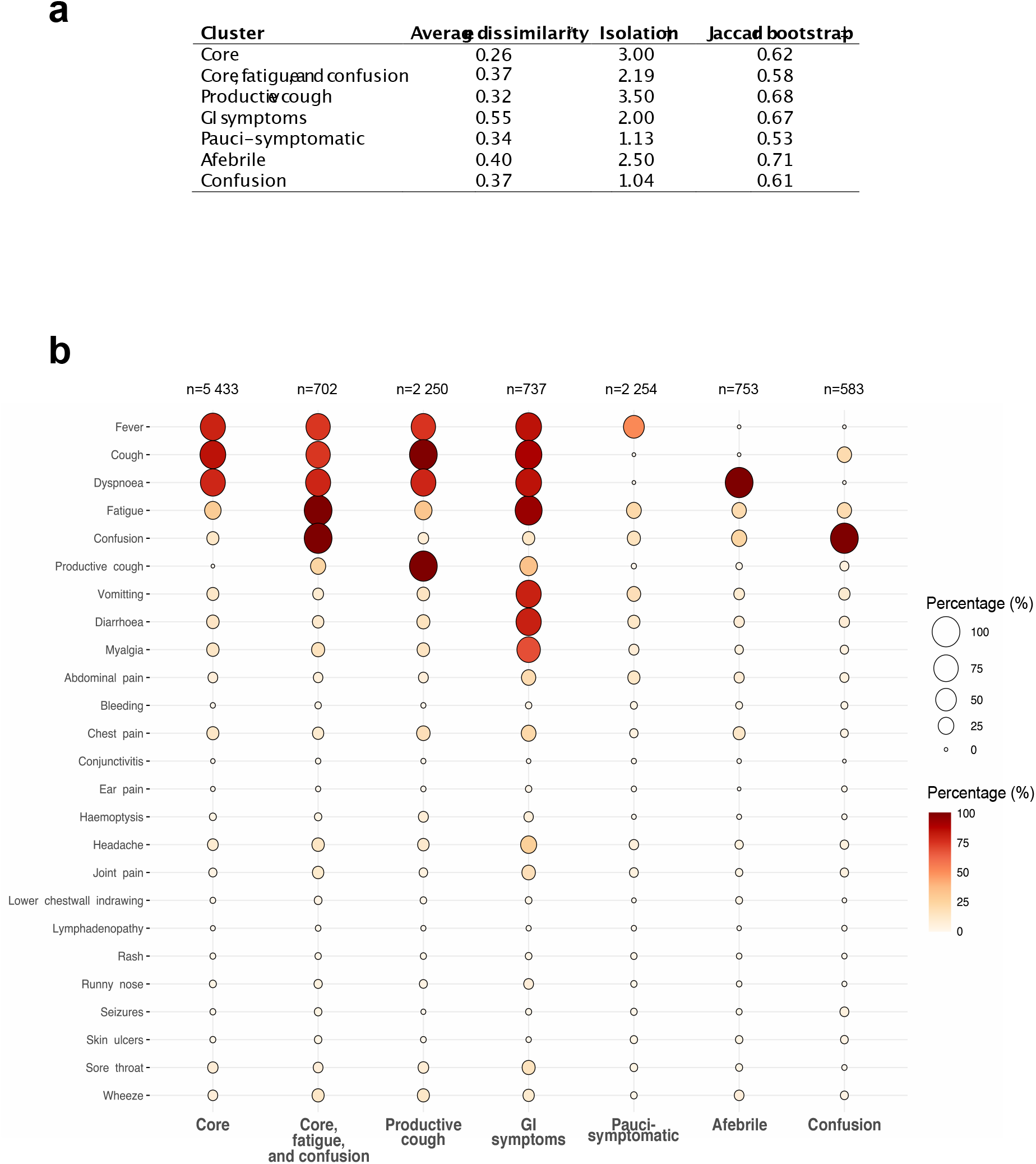
Internal validity, and sensitivity analysis. a. Meaures of clustering internal validity and stability for k=7. \*Average dissimilarity - average dissimilarity between observations in the cluster and the cluster medoid. †Isolation maximal disimilarity between observations in the cluster and the cluster medoid/minimal disimilarity between the cluster medoid and the medoid of any other cluster. A smaller ratio suggests the cluster is well isolated. ‡Jaccard bootstrap - the clusterwise Jaccard coefficient for 1000 iterations of the clustering algorithm on random subsets of the data. **c**. Sensitivity analysis. Resultant clusters and patterns of symptoms in cases with fully complete symptom data (n=12712). Data are presented as count (percentage).

**Supplementary Figure S5.**
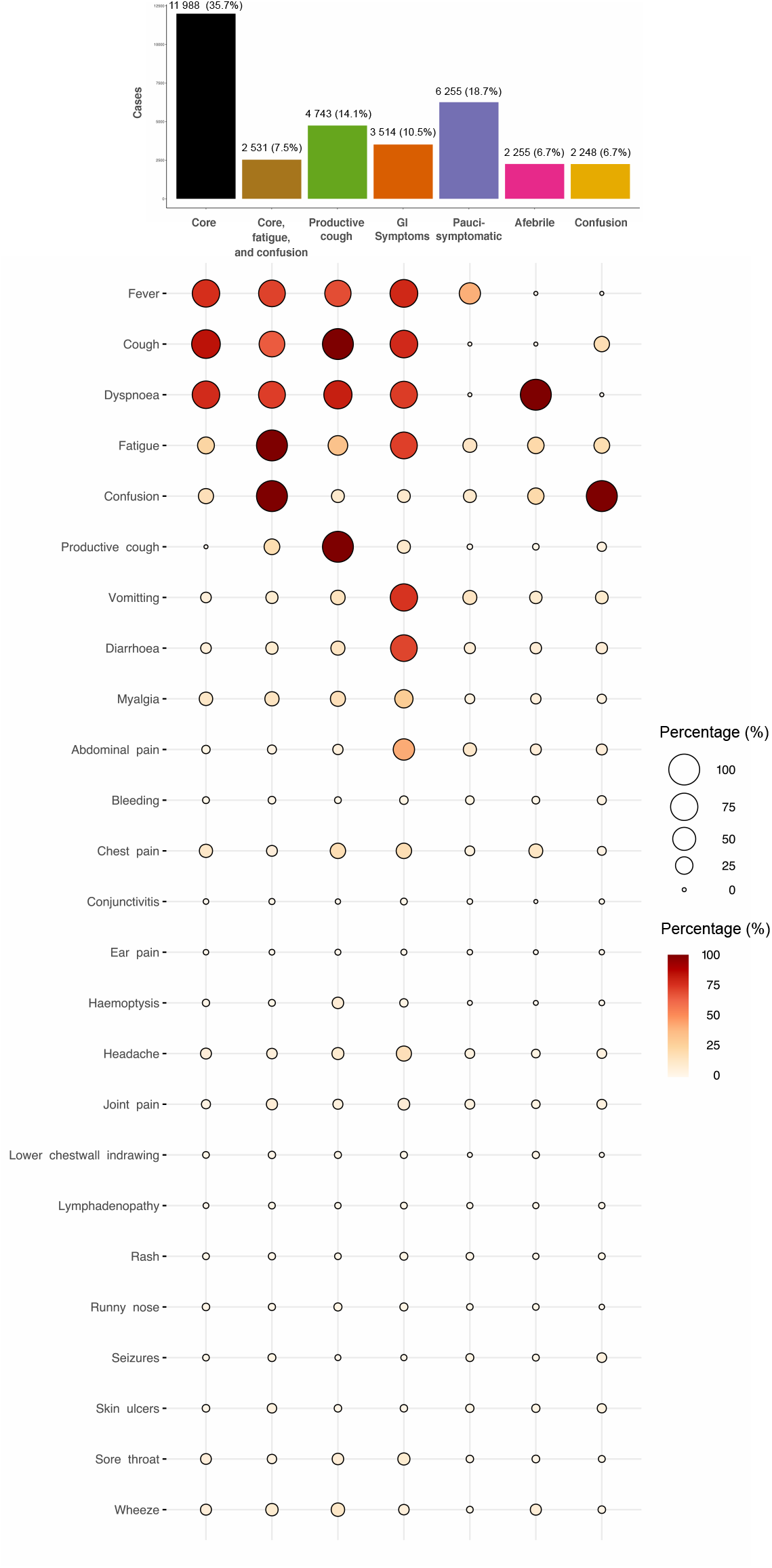
Internal clustering validation. Cluster identities, proportions, and patterns in patients enrolled to ISARIC CCP-UK after the primary analysis until 7th July, 2020 (n=33 534). Data are presented as Data are presented as count (percentage).

**Supplementary Figure 6.**
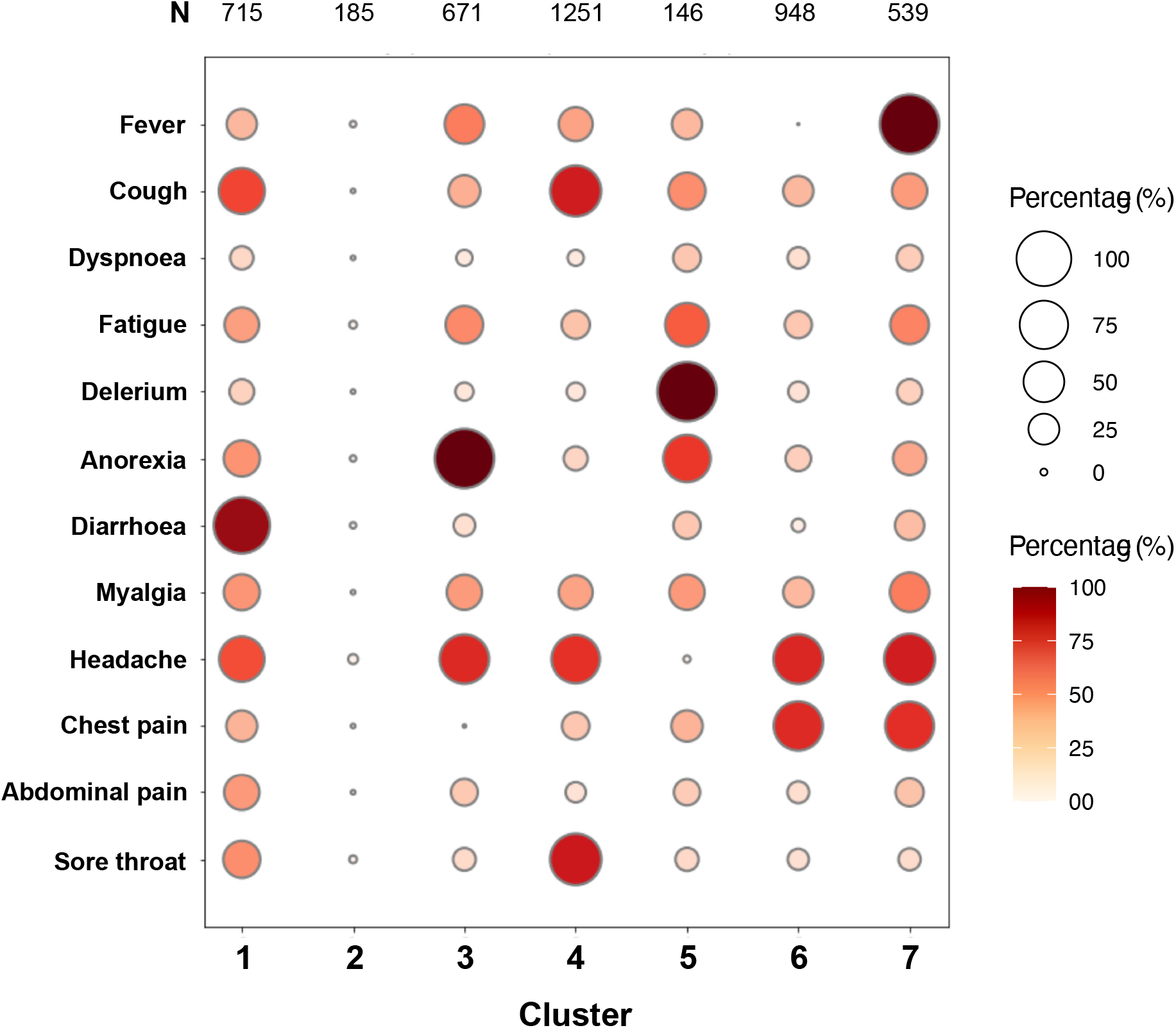
Clustering validation in the COVID Symptom Study dataset. Symptomatic individuals with laboratory confirmed SARS-CoV-2 infection, and who logged symptoms more than twice within the first week of onset. Data are presented as percentage of patients within a cluster exhibiting a given symptom.

**Supplementary Table 1.** Patient characteristics and outcomes.

|  | N | Core<br>n = 9364 | Core,<br>fatigue,<br>and confusion<br>n = 1796 | Productive<br>cough<br>n = 4234 | GI symptoms<br>n = 1327 | Pauci-symptomatic<br>n = 3692 | Afebrile<br>n = 3624 | Confusion<br>n = 1440 | Overall<br>n = 25477 |
| --- | --- | --- | --- | --- | --- | --- | --- | --- | --- |
| **Age** | 23493 | 68 (55, 82) | 69 (55, 81) | 72 (58, 81) | 60 (49, 72) | 76 (63, 85) | 76 (64, 85) | 82 (75, 88) | 73 (59, 83) |
| **Sex (male)** | 25359 | 5735 (61%) | 1106 (62%) | 2600 (62%) | 699 (53%) | 2033 (55%) | 2084 (58%) | 789 (55%) | 15,046 (59%) |
| **Ethnicity** | 22643 |  |  |  |  |  |  |  |  |
| Aboriginal/First Nations |  | 2 (<0.1%) | 0 (0%) | 1 (<0.1%) | 0 (0%) | 1 (<0.1%) | 2 (<0.1%) | 1 (<0.1%) | 7 (<0.1%) |
| Arab |  | 55 (1%) | 6 (<1%) | 19 (1%) | 4 (<1%) | 14 (<1%) | 16 (1%) | 3 (<1%) | 117 (1%) |
| Black |  | 351 (4%) | 46 (3%) | 163 (4%) | 40 (3%) | 119 (4%) | 87 (3%) | 31 (2%) | 837 (4%) |
| East Asian |  | 100 (1%) | 16 (1%) | 24 (1%) | 13 (1%) | 29 (1%) | 23 (1%) | 6 (1%) | 211 (1%) |
| South Asian |  | 412 (5%) | 50 (3%) | 167 (4%) | 71 (6%) | 102 (3%) | 116 (4%) | 18 (1%) | 936 (4%) |
| West Asian |  | 36 (<1%) | 3 (<1%) | 14 (<1%) | 5 (<1%) | 11 (<1%) | 12 (<1%) | 7 (1%) | 88 (<1%) |
| Latin American |  | 17 (<1%) | 1 (<0.1%) | 9 (<1%) | 2 (<1%) | 7 (<1%) | 5 (<1%) | 1 (<0.1%) | 42 (<1%) |
| White |  | 6592 (80%) | 1441 (81%) | 3100 (82%) | 960 (81%) | 2807 (85%) | 2578 (86%) | 1166 (91%) | 18,824 (83%) |
| Other |  | 687 (8%) | 94 (6%) | 266 (7%) | 91 (8%) | 194 (6%) | 194 (6%) | 55 (4%) | 1581 (7%) |
| **Co-morbidities** |  |  |  |  |  |  |  |  |  |
| Chronic cardiac disease | 23933 | 2243 (25%) | 648 (38%) | 1289 (32%) | 244 (19%) | 1090 (32%) | 1302 (39%) | 602 (44%) | 7418 (31%) |
| Diabetes | 23670 | 1701 (20%) | 395 (23%) | 724 (18%) | 238 (19%) | 625 (18%) | 723 (22%) | 266 (20%) | 4672 (20%) |
| Obesity | 21707 | 913 (11%) | 146 (9%) | 480 (13%) | 193 (16%) | 200 (6%) | 291 (10%) | 65 (5%) | 2288 (11%) |
| Malnutrition | 22511 | 119 (1%) | 69 (4%) | 67 (2%) | 21 (2%) | 105 (3%) | 79 (3%) | 66 (5%) | 526 (2%) |
| Chronic pulmonary disease, not<br>asthma | 23828 | 1229 (14%) | 351 (21%) | 1118 (28%) | 128 (10%) | 477 (14%) | 780 (24%) | 223 (17%) | 4306 (18%) |
| Asthma | 23732 | 1279 (15%) | 177 (10%) | 812 (20%) | 205 (16%) | 297 (9%) | 444 (14%) | 129 (10%) | 3343 (14%) |
| Chronic renal disease | 23685 | 1238 (14%) | 342 (20%) | 571 (14%) | 126 (10%) | 653 (19%) | 610 (19%) | 310 (23%) | 3850 (16%) |
| Moderate/severe liver disease | 23481 | 120 (1%) | 41 (2%) | 57 (1%) | 18 (1%) | 69 (2%) | 69 (2%) | 41 (3%) | 415 (2%) |
| Mild liver disease | 23431 | 107 (1%) | 39 (2%) | 73 (2%) | 25 (2%) | 41 (1%) | 59 (2%) | 16 (1%) | 360 (2%) |
| Chronic neurological disease | 23530 | 862 (10%) | 327 (9%) | 340 (9%) | 55 (4%) | 504 (15%) | 355 (11%) | 278 (21%) | 2721 (12%) |
| Dementia | 23630 | 1050 (12%) | 447 (26%) | 271 (7%) | 23 (2%) | 650 (20%) | 466 (14%) | 467 (35%) | 3374 (14%) |
| Chronic rheumatological disease | 23407 | 783 (9%) | 182 (11%) | 403 (10%) | 123 (10%) | 347 (10%) | 332 (10%) | 167 (13%) | 2337 (10%) |
| Chronic haematological disease | 23454 | 340 (4.0%) | 76 (5%) | 169 (4%) | 35 (3%) | 163 (5%) | 129 (4%) | 52 (4%) | 964 (4%) |
| Malignancy | 23487 | 701 (8%) | 196 (12%) | 422 (11%) | 90 (7%) | 410 (12%) | 343 (11%) | 158 (12%) | 2320 (10%) |
| HIV | 23311 | 37 (<1%) | 7 (<1%) | 22 (1%) | 10 (1%) | 10 (<1%) | 15 (1%) | 3 (<1%) | 104 (%) |
| **Smoking status** | 18357 |  |  |  |  |  |  |  |  |
| Former Smoker |  | 1935 (29%) | 469 (36%) | 1232 (37%) | 345 (32%) | 690 (28%) | 859 (34%) | 308 (33%) | 5838 (32%) |
| Never Smoked |  | 4396 (66%) | 735 (57%) | 1879 (56%) | 702 (64%) | 1603 (64%) | 1432 (58%) | 556 (60%) | 11,321 (62%) |
| Yes |  | 362 (5%) | 85 (7%) | 235 (7%) | 44 (4%) | 207 (8%) | 195 (8%) | 70 (8%) | 1198 (6.5%) |
| **Outcomes** |  |  |  |  |  |  |  |  |  |
| ICU admission | 24443 | 1841 (21%) | 224 (13%) | 700 (17%) | 293 (23%) | 266 (7.5%) | 453 (13%) | 69 (5.0%) | 3846 (16%) |
| Mechanical ventilation | 23907 | 1073 (12%) | 468 (13%) | 137 (8%) | 153 (12%) | 130 (4%) | 268 (8%) | 35 (3%) | 2174 (9%) |
| Mortality | 19358 | 2402 (33%) | 741 (53%) | 1104 (33%) | 186 (18%) | 830 (31%) | 1086 (40%) | 502 (48%) | 6851 (35%) |
| Unknown Mortality | 6119 | 2191 (23%) | 389 (22%) | 889 (21%) | 297 (22%) | 1029 (28%) | 920 (25%) | 404 (28%) | 6119 (24%) |

**Supplementary Table 2.** Symptom prevalence.

| **Symptom** | **Number of patients (n=25 477)** | **Prevalence (95% CI)** |
| --- | --- | --- |
| Cough | 17 334 | 68.0 (67.5-68.6) |
| Fever | 16 920 | 66.4 (65.8-67.0) |
| Dyspnoea | 16 603 | 65.2 (64.6-65.8) |
| Fatigue | 8 906 | 35.0 (34.4-35.5) |
| Confusion | 5 935 | 23.3 (22.8-23.8) |
| Productive cough | 5 125 | 20.1 (19.6-20.6) |
| Diarrhoea | 4 171 | 16.4 (15.9-16.8) |
| Vomiting | 4 144 | 16.3 (15.8-16.7) |
| Myalgia | 3 719 | 14.6 (14.2-15.0) |
| Chest pain | 2 925 | 11.5 (11.1-11.9) |
| Headache | 2 213 | 8.7 (8.3-9.0) |
| Abdominal pain | 2 094 | 8.2 (7.9-8.6) |
| Wheeze | 2 029 | 8.0 (7.6-8.3) |
| Sore throat | 1 731 | 6.8 (6.5-7.1) |
| Joint pain | 1 294 | 5.1 (4.8-5.4) |
| Haemoptysis | 652 | 2.6 (2.4-2.8) |
| Runny nose | 583 | 2.3 (2.1-2.5) |
| Skin ulcers | 473 | 1.9 (1.7-2.0) |
| Seizures | 336 | 1.3 (1.2-1.5) |
| Rash | 307 | 1.2 (1.1-1.3) |
| Bleeding | 259 | 1.0 (0.9-1.1) |
| Chest wall in-drawing | 254 | 1.0 (0.9-1.1) |
| Lymphadenopathy | 119 | 0.5 (0.4-0.6) |
| Ear pain | 98 | 0.4 (0.3-0.5) |
| Conjunctivitis | 73 | 0.3 (0.2-0.4) |

**Supplementary Table 3.** Multinomial regression model odds ratios.

|  | Cluster | Relative risk ratio (RRR) | 95% CI | P value |
| --- | --- | --- | --- | --- |
| Male sex | Core | *reference* |  |  |
|  | Core, fatigue, and cough | 1.02 | 0.91-1.14 | 0.743 |
|  | Productive cough | 1.02 | 0.94-1.10 | 0.660 |
|  | GI symptoms | 0.65 | 0.57-0.73 | <0.001 |
|  | Pauci-symptomatic | 0.80 | 0.73-0.87 | <0.001 |
|  | Afebrile | 0.85 | 0.78-0.92 | <0.001 |
|  | Confusion | 0.81 | 0.72-0.91 | <0.001 |
| Age 0-20 | Core | *reference* |  |  |
|  | Core, fatigue, and cough | 0.28 | 0.12-0.64 | 0.002 |
|  | Productive cough | 0.46 | 0.30-0.72 | <0.001 |
|  | GI symptoms | 0.74 | 0.42-1.30 | 0.295 |
|  | Pauci-symptomatic | 3.59 | 2.81-4.57 | <0.001 |
|  | Afebrile | 0.97 | 0.68-1.37 | 0.863 |
|  | Confusion | 0.55 | 0.26-1.13 | 0.101 |
| Age 20-40 | Core | *reference* |  |  |
|  | Core, fatigue, and cough | 0.35 | 0.24-0.51 | <0.001 |
|  | Productive cough | 0.97 | 0.82-1.16 | 0.771 |
|  | GI symptoms | 1.34 | 1.06-1.70 | 0.014 |
|  | Pauci-symptomatic | 0.89 | 0.74-1.07 | 0.221 |
|  | Afebrile | 0.66 | 0.54-0.81 | <0.001 |
|  | Confusion | 0.20 | 0.11-0.36 | <0.001 |
| Age 40-60 | Core | *reference* |  |  |
|  | Core, fatigue, and cough | 0.38 | 0.32-0.46 | <0.001 |
|  | Productive cough | 0.83 | 0.75-0.93 | <0.001 |
|  | GI symptoms | 1.30 | 1.12-1.51 | <0.001 |
|  | Pauci-symptomatic | 0.48 | 0.42-0.55 | <0.001 |
|  | Afebrile | 0.56 | 0.50-0.64 | <0.001 |
|  | Confusion | 0.27 | 0.21-0.35 | <0.001 |
| Age >80 | Core | *reference* |  |  |
|  | Core, fatigue, and cough | 1.46 | 1.30-1.66 | <0.001 |
|  | Productive cough | 1.08 | 0.98-1.19 | 0.123 |
|  | GI symptoms | 0.53 | 0.44-0.65 | <0.001 |
|  | Pauci-symptomatic | 1.46 | 1.33-1.61 | <0.001 |
|  | Afebrile | 1.38 | 1.26-1.52 | <0.001 |
|  | Confusion | 2.11 | 1.85-2.41 | <0.001 |
| Chronic cardiac disease | Core | *reference* |  |  |
|  | Core, fatigue, and cough | 1.13 | 1.00-1.27 | 0.041 |
|  | Productive cough | 1.24 | 1.13-1.36 | <0.001 |
|  | GI symptoms | 1.01 | 0.86-1.20 | 0.868 |
|  | Pauci-symptomatic | 1.03 | 0.93-1.13 | 0.569 |
|  | Afebrile | 1.35 | 1.23-1.48 | <0.001 |
|  | Confusion | 1.36 | 1.20-1.54 | <0.001 |
| Chronic pulmonary disease | Core | *reference* |  |  |
|  | Core, fatigue, and cough | 1.25 | 1.09-1.43 | 0.002 |
|  | Productive cough | 2.17 | 1.97-2.39 | <0.001 |
|  | GI symptoms | 0.81 | 0.66-1.00 | 0.054 |
|  | Pauci-symptomatic | 0.84 | 0.74-0.94 | 0.004 |
|  | Afebrile | 1.48 | 1.33-1.65 | <0.001 |
|  | Confusion | 0.87 | 0.74-1.03 | 0.105 |
| Asthma | Core | *reference* |  |  |
|  | Core, fatigue, and cough | 0.82 | 0.69-0.98 | 0.028 |
|  | Productive cough | 1.50 | 1.35-1.67 | <0.001 |
|  | GI symptoms | 0.99 | 0.81-1.18 | 0.915 |
|  | Pauci-symptomatic | 0.62 | 0.54-0.72 | <0.001 |
|  | Afebrile | 0.94 | 0.83-1.06 | 0.334 |
|  | Confusion | 0.78 | 0.64-0.95 | 0.013 |
| Chronic kidney disease | Core | *reference* |  |  |
|  | Core, fatigue, and cough | 1.05 | 0.91-1.21 | 0.509 |
|  | Productive cough | 0.87 | 0.78-0.98 | 0.023 |
|  | GI symptoms | 0.90 | 0.73-1.11 | 0.344 |
|  | Pauci-symptomatic | 1.18 | 1.06-1.32 | 0.003 |
|  | Afebrile | 1.01 | 0.90-1.13 | 0.883 |
|  | Confusion | 1.09 | 0.94-1.26 | 0.276 |
| Chronic neurological disease | Core | *reference* |  |  |
|  | Core, fatigue, and cough | 1.69 | 1.45-1.95 | <0.001 |
|  | Productive cough | 0.92 | 0.80-1.06 | 0.238 |
|  | GI symptoms | 0.51 | 0.38-0.68 | <0.001 |
|  | Pauci-symptomatic | 1.37 | 1.21-1.56 | <0.001 |
|  | Afebrile | 1.01 | 0.88-1.15 | 0.940 |
|  | Confusion | 1.76 | 1.50-2.06 | <0.001 |
| Malignancy | Core | *reference* |  |  |
|  | Core, fatigue, and cough | 1.21 | 1.01-1.44 | 0.035 |
|  | Productive cough | 1.28 | 1.11-1.46 | <0.001 |
|  | GI symptoms | 1.13 | 0.89-1.44 | 0.324 |
|  | Pauci-symptomatic | 1.35 | 1.18-1.55 | <0.001 |
|  | Afebrile | 1.13 | 0.98-1.30 | 0.095 |
|  | Confusion | 1.14 | 0.94-1.38 | 0.183 |
| Chronic haematological disease | Core | *reference* |  |  |
|  | Core, fatigue, and cough | 0.99 | 0.76-1.30 | 0.962 |
|  | Productive cough | 1.01 | 0.83-1.24 | 0.885 |
|  | GI symptoms | 0.72 | 0.49-1.05 | 0.088 |
|  | Pauci-symptomatic | 1.00 | 0.82-1.23 | 0.975 |
|  | Afebrile | 0.84 | 0.68-1.05 | 0.125 |
|  | Confusion | 0.83 | 0.61-1.12 | 0.223 |
| HIV | Core | *reference* |  |  |
|  | Core, fatigue, and cough | 1.03 | 0.42-2.57 | 0.944 |
|  | Productive cough | 1.58 | 0.91-2.73 | 0.103 |
|  | GI symptoms | 1.97 | 0.96-4.02 | 0.063 |
|  | Pauci-symptomatic | 0.95 | 0.46-1.95 | 0.887 |
|  | Afebrile | 1.40 | 0.75-2.60 | 0.291 |
|  | Confusion | 0.25 | 0.03-1.86 | 0.175 |
| Chronic rheumatological disease | Core | *reference* |  |  |
|  | Core, fatigue, and cough | 0.99 | 0.83-1.19 | 0.953 |
|  | Productive cough | 1.05 | 0.91-1.20 | 0.509 |
|  | GI symptoms | 1.24 | 1.00-1.54 | 0.053 |
|  | Pauci-symptomatic | 0.97 | 0.84-1.11 | 0.622 |
|  | Afebrile | 0.92 | 0.80-1.06 | 0.256 |
|  | Confusion | 1.02 | 0.85-1.23 | 0.804 |
| Dementia | Core | *reference* |  |  |
|  | Core, fatigue, and cough | 1.58 | 1.38-1.82 | <0.001 |
|  | Productive cough | 0.48 | 0.42-0.56 | <0.001 |
|  | GI symptoms | 0.22 | 0.15-0.34 | <0.001 |
|  | Pauci-symptomatic | 1.15 | 1.02-1.29 | 0.089 |
|  | Afebrile | 0.81 | 0.72-0.92 | 0.001 |
|  | Confusion | 1.80 | 1.57-2.07 | <0.001 |
| Malnutrition | Core |  | *reference* |  |
|  | Core, fatigue, and cough | 2.20 | 1.61-3.01 | <0.001 |
|  | Productive cough | 1.21 | 0.89-1.66 | 0.225 |
|  | GI symptoms | 1.39 | 0.82-2.33 | 0.220 |
|  | Pauci-symptomatic | 1.77 | 1.34-2.34 | <0.001 |
|  | Afebrile | 1.47 | 1.10-1.97 | 0.012 |
|  | Confusion | 2.45 | 1.78-3.38 | <0.001 |
| Chronic liver disease | Core |  | *reference* |  |
|  | Core, fatigue, and cough | 2.09 | 1.58-2.77 | <0.001 |
|  | Productive cough | 1.20 | 0.94-1.52 | 0.142 |
|  | GI symptoms | 1.20 | 0.83-1.73 | 0.338 |
|  | Pauci-symptomatic | 1.35 | 1.05-1.75 | 0.019 |
|  | Afebrile | 1.61 | 1.27-2.04 | <0.001 |
|  | Confusion | 2.16 | 1.57-2.97 | <0.001 |
| Diabetes | Core |  | *reference* |  |
|  | Core, fatigue, and cough | 1.20 | 1.06-1.35 | <0.001 |
|  | Productive cough | 0.89 | 0.81-0.98 | 0.014 |
|  | GI symptoms | 1.12 | 0.97-1.30 | 0.123 |
|  | Pauci-symptomatic | 1.00 | 0.91-1.10 | 0.988 |
|  | Afebrile | 1.08 | 0.98-1.19 | 0.108 |
|  | Confusion | 1.09 | 0.95-1.24 | <0.001 |

**Supplementary Table 4.** Restricted mean survival times adjusted for age.

| Cluster | Sex | Mean difference (days) | 95% CI (days) | P value |
| --- | --- | --- | --- | --- |
| Core |  | *Reference cluster* |  |  |
| Core + fatigue + confusion | Male | -1.9 | -2.8 – -1.1 | <0.001 |
|  | Female | -1.7 | -2.8 – -0.6 | 0.002 |
| Productive cough | Male | 0.5 | -0.1 – 1.0 | 0.085 |
|  | Female | 0.5 | -0.2 – 1.2 | 0.132 |
| GI symptoms | Male | 1.3 | 0.5 – 2.1 | 0.002 |
|  | Female | 2.7 | 2.0 – 3.5 | <0.001 |
| Pauci-symptomatic | Male | 2.5 | 1.9 – 3.0 | <0.001 |
|  | Female | 2.3 | 1.7 – 2.9 | <0.001 |
| Afebrile | Male | 0.4 | -0.3 – 1.0 | 0.258 |
|  | Female | 0.3 | -0.4 – 1.0 | 0.426 |
| Confusion | Male | 0.7 | -0.3 – 1.7 | 0.158 |
|  | Female | 1.2 | 0.1 – 2.3 | 0.038 |

